# Development of an extraction-free LAMP method for the generic detection of Orthopoxvirus and for the specific detection of Mpox virus

**DOI:** 10.1101/2023.06.21.23291715

**Authors:** Zhiru Li, Amit Sinha, Yinhua Zhang, Nathan Tanner, Hui-Ting Cheng, Prem Premsrirut, Clotilde K. S. Carlow

**Affiliations:** New England Biolabs, Ipswich, Massachusetts, 01938, USA; Mirimus Inc. Brooklyn, New York, 11226, USA

**Keywords:** Orthopoxvirus, Monkeypox, Mpox virus, LAMP, Extraction free

## Abstract

Mpox is a neglected zoonotic disease endemic in West and Central Africa. The 2022 Mpox outbreak with more than 18,000 cases worldwide generated great concern about future outbreaks and highlighted the need for a simple and rapid diagnostic test. The Mpox virus, MPXV, is a member of the Orthopoxvirus genus that also contains other pathogenic viruses including variola virus, vaccinia virus, camelpox virus, and cowpox virus. Phylogenomic analysis of all available Orthopoxvirus genomes identified 10 distinct phylogroups, with isolates from infected humans distributed across various phylogroups interspersed with isolates from animal sources, indicating the zoonotic potential of these viruses. In this study, we developed a simple and sensitive colorimetric pan-Orthopoxvirus LAMP assay for broader Orthopoxvirus detection. We also developed an MPXV-specific probe that differentiates MPXV from other Orthopoxviruses on the N1R gene which differs by only a few nucleotides. In addition, we described an extraction-free protocol for use directly with swab eluates in LAMP assays, thereby eliminating the time and resources needed to extract DNA from the sample. Our direct LAMP assays are well-suited for low-resource settings and provide a valuable tool for rapid and scalable diagnosis and surveillance of Orthopoxviruses and MPXV.

## Introduction

Mpox (previously known as Monkeypox) was first described in 1958 in laboratory monkeys shipped from Singapore to Denmark [1]. The first Mpox case in humans was reported in 1970 in the Democratic Republic of the Congo [2] followed by sporadic outbreaks mainly in West and Central Africa [3]. Even though about 400 confirmed cases of Mpox from these areas were reported in the last two decades, the suspected number of cases has been predicted to be much higher, to around 28,000 [4]. While Mpox remains a neglected tropical disease, the 2003 outbreak in the United States generated international attention and the global outbreak in 2022 with more than 85,000 cases in more than 100 countries led to the declaration of a Public Health Emergency of International Concern by the World Health Organization [5]. Mpox is caused by the Mpox virus (MPXV), which is an enveloped double-stranded DNA virus with an AT-rich (67% on average) genome of about 200 kb encoding 190 open-reading frames [6]. It is a member of the Orthopoxvirus genus that also contains several viruses of great medical relevance, including variola virus, the causative agent of smallpox; vaccinia virus, the virus used in the smallpox vaccine; cowpox, camelpox, and a few other species that could infect humans [7]. Currently, MPXV is classified into two major clades, each with distinct geographical, clinical, genomic, and epidemiological differences [8]. Viruses from Clade I (former Congo Basin clade) cause more clinically severe disease in humans, with higher mortality rates and transmissibility, while those from Clade IIa (former West African clade) have a milder clinical presentation with lower mortality rates and transmissibility. The isolates from the most recent 2022 outbreak are placed in Clade IIb and closely related to the virus responsible for the 2017-2019 outbreaks in the same clade [8–10].

Human-to-human transmission of MPXV can occur by direct contact, respiratory secretions, vertical transmission, or indirect contact through fomites. Direct contact with infectious sores or lesions on mucous membranes has been the primary mode of transmission during the 2022 outbreak [11]. Mpox diagnosis is based on suspected epidemiological and clinical symptoms and confirmed by nucleic acid amplification testing, mainly real-time PCR. The recommended diagnostic specimens are directly from skin lesions or biopsies [12, 13]. Viral DNA can also be detected in saliva [14], semen, blood, or urine samples, even though usually less frequently and with a lower viral load [15–17] but could potentially help early detection of Mpox before the development of skin lesions. At present, detecting viral DNA by quantitative polymerase chain reaction (qPCR) is the recommended laboratory test for Mpox [18]; indeed, a positive PCR result is considered definitive, regardless of associated symptoms. However, the qPCR method requires complex and expensive equipment. In rural areas or low-resource settings without access to high-precision PCR instruments, a faster and simpler method for Mpox testing is needed as a viable alternative. Loop-mediated isothermal amplification (LAMP) is a nucleic acid amplification method that uses a Bst DNA polymerase with strand displacement activity. The assay is conducted under isothermal conditions ranging from 60-66°C and is fast and simple to use, making it ideal for the diagnosis and surveillance of neglected tropical diseases [19]. LAMP diagnostic assays have also been successfully used as a point-of-care diagnostic tool during the SARS-CoV-2 pandemic [20–22].

In this study, we developed a pan-Orthopoxvirus colorimetric LAMP assay as well as a fluorescent probe-based MPXV-specific LAMP assay. In addition, we described a protocol to detect viral DNA directly from the swab samples without the need for a DNA extraction step. Given the constant threat and continued transmission of MPXV in different parts of the world [23], accessibility to testing is critical for worldwide control efforts. Our simple and quick colorimetric LAMP assay can improve the testing capability, especially in remote and low-resource areas.

## Materials and methods

### Ethical approval

All samples submitted in the study were obtained in accordance with ethical guidelines and remain anonymous. Any identifying information was de-identified to maintain confidentiality and the samples were labeled using a barcode for identification purposes only. The de-identified (barcoded) samples (swabs from lesions) were submitted to Mirimus Clinical Laboratory and collected within the interim guidelines for laboratory testing established by the WHO (May 23^rd^, 2022). Sample collection and submission methods for use in the Mirimus Clinical Laboratory study protocol were reviewed and approved by Advarra Institutional Review Board Pro00065623.

### Phylogenomic and homolog analysis

Genome assemblies for all OPXV available as on May 18^th^, 2022 were downloaded from GenBank, and protein coding genes in each assembly were annotated using PROKKA version 1.14.6 [24]. Genomes of Centapox viruses Yokapox, Murmansk virus and Centapox NY014 were downloaded for use as outgroups. Metadata on the source of isolation of each virus was compiled (S1 Table). Orthology analysis and identification of single copy orthologs for phylogenomic analysis were performed using Orthofinder version 2.4.0 [25]. The gene sequences corresponding to the identified SCOs were concatenated together to generate a supermatrix sequence for each species, and their multiple sequence alignment was obtained using mafft v7.149b [26]. Phylogenetic analysis of the resulting supermatrix of 33,425 nucleotides was carried out using W-IQ-TREE [27] where the best-fit substitution model was chosen automatically using ModelFinder [28] and bootstrap support values were calculated based on ultrafast bootstrap [29] with 1000 replicates. The OPXV clades were named as phylogroups 1 to 10 and denoted as OPXV-PG-1 to OPXV-PG-10. To identify orthogroups that are present or absent across various phylogroups, protein sequences from each isolate within a phylogroup were first combined into a pan-phylogroup proteome and were then used for orthology analysis using OrthoFinder version 2.4.0 [25]. The conservation of orthogroups across various phylogroups was visualized as an UpSet plot [30]. Homologs of MPXV A4L and N1R genes were identified through NCBI Blast search. Alignments of downloaded homologs were aligned using ClustalW (https://www.genome.jp/tools-bin/clustalw).

### Control virus DNA and MPXV samples

Genomic DNA from MPXV USA-2003 (BEI, #NR-4928, Lot 70053399, at least 98% identical to GenBank accession number NC_063383) and synthetic hMPXV control 2 (Twist Bioscience, # 106059, Lot 2000006347 identical to GenBank accession number NC_063383) were used as the template for LAMP assays. Genomic DNA from camelpox virus strain V78-2379 (BEI, #NR-50076, Lot 64108452, 99% identical to GenBank accession number NC_003391) and vaccinia virus WR (BEI #NR-2640, Lot 60981371, GenBank: NC_006998) were also used for specificity analysis. Three clinical MPXV DNA samples and their corresponding swab eluates samples were donated by Mirimus Inc. Human genomic DNA (Promega #G3041, Lot 0000531679) was used as the negative control. Viral DNA was aliquoted into small volumes and stored at −80°C until use.

### Real-Time PCR

Real-Time PCR was performed using the Luna® Universal Probe qPCR Master (NEB #M3004) following the manufacturer’s instructions. Each 20 μL reaction contained 2 μL template DNA. CDC-Non-Variola Orthopoxvirus Forward primer (5’-TCAACTGAAAAGGCCATCTAT GA-3’), Reverse primer (5’-GAGTATAGAGCACTATTTCTAAATCCCA-3’) and dual quencher modified probe (5’-FAM-CCATGCAAT/ZEN/ATACGTACAAGATAGTAGCCAAC-3’IABkFQ) were used [31]. The reactions were performed on a Bio-Rad CFX Opus instrument using the following cycling conditions: initial denaturation (95°C for 1 min) followed by 45 cycles of alternating denaturation (95°C for 10 sec) with annealing/elongation (60°C for 30 sec) plus a plate read step.

### Colorimetric and probe-based LAMP assay

LAMP primer design was based on MPXV reference genomes NC_003310 and NC_063383. Multiple genes were chosen for LAMP primer design using the NEB Primer Design Tool (https://lamp.neb.com/). Among the 14 sets of LAMP primers (S2 Table) tested targeting various genes, 2 sets of primers showed the best performance and were chosen for this study (Table 1). Each primer set included an outer forward primer (F3), outer backward primer (B3), forward inner primer (FIP), backward inner primer (BIP), forward loop primer (LF), and backward loop primer (LB). Primers were synthesized by Integrated DNA Technologies™ (Coralville, IA, USA).

**Table 1.**
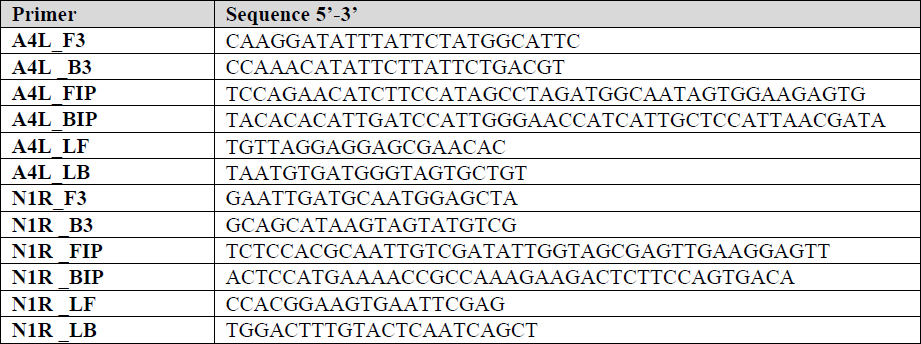
LAMP primer sequences targeting A4L and N1R.

The colorimetric LAMP assay was performed using the WarmStart® Colorimetric LAMP 2X Master Mix with UDG (NEB #M1804). Each 20 μL reaction contained 10 µL 2X Master Mix, 2 µL 10X primer mix to their final concentration F3/B3 0.2 µM each; FIP/BIP 1.6 µM each; LF/LB 0.4 µM each), 2 µL 10X guanidine hydrochloride (400 mM), 2-4 µL of DNA or crude swab eluate, and DNase/RNase free water. Reactions were assembled in 96-well plates on ice followed by incubation at 65°C for up to 1 hour. Samples were considered positive for the presence of the virus if the reaction had a color change from pink to yellow, or negative if the reaction remained pink. To record color changes, 96-well plates were imaged using an Epson Perfection V600 Photo Scanner before and after the LAMP reaction. To enable reaction dynamics to be monitored in real-time, 1 µM of SYTO™ 9 Green Fluorescent Nucleic Acid Stain (Invitrogen) was included in the LAMP reaction and the reactions performed in a qPCR machine (Bio-Rad CFX Opus). The Cq number was converted to Tt which represents the time in minutes to reach the fluorescence detection threshold, using a conversion factor of 22 seconds per cycle. No amplification is denoted N/A. Experiments were conducted using at least two replicates.

Probe-based LAMP assays were performed with a fluorescent-based LAMP kit (NEB #E1700) with 0.25µM MPXV_Probe (/5Cy5/T+T+GTGCAA+T+AAT+TGGAC/3IAbRQSp/) instead of the fluorescent dye included in the kit. The hybridization probe contains a 5′ Cy5 fluorophore and a 3’ end dark quencher and locked nucleic acid (LNA) bases indicated by a preceding + symbol. The probe was synthesized by Integrated DNA Technologies™ (Coralville, IA, USA). All probe-based reactions were performed in a qPCR machine (Bio-Rad CFX Opus).

### Molecular testing of clinical swab samples

De-identified (barcoded) swab samples were used within the study. Dry swab specimens were rehydrated in 1 mL TE buffer (ThermoFisher, Catalog#J75793-AP). For the standard qPCR test, 100 µL of eluate was subjected to nucleic acid extraction and purification using the KingFisher Flex automated nucleic acid extraction (ThermoFisher, Catalog A48383), and 2.5 µL of purified DNA was amplified and detected using 4X Luna MasterMIx (NEB, Catalog#M3019B) and CDC-defined primer/probe sets (S2 Table) in a total of 5 µL of reaction. For extraction-free LAMP test, swab eluates were treated similarly as saliva as previously described [20]. Briefly, an equal volume of eluates was mixed with an equal volume of 2X SLB buffer containing 5 mM tris(2-carboxyethyl) phosphine (TCEP, Millipore Cat# 580567), 22 mM sodium hydroxide (Sigma 72068), 2 mM Ethylenediaminetetraacetic acid (EDTA, Invitrogen 15575–038) and 0.4% Pluronic F-68 (Gibco 24040–032). Mixed samples were heated in a thermocycler at 95°C for 5 minutes, then cooled to 4°C. For each LAMP reaction, 4 μL of the treated sample, corresponding to 2 μL of swab eluate, was used in the 20 μL reaction.

## Results

### Phylogenomic analysis of Orthopoxvirus genomes

A phylogenomic analysis of 200 Orthopoxvirus genomes, identified 10 distinct clades within this genus, which are here named as phylogroups 1 to 10 and are denoted as OPXV-PG-01 to OPXV-PG-10 (Fig 1) based on their nesting patterns (S1 Fig), where the 100 MPXV isolates included in this analysis are placed in OPXV-PG-06. Two distinct clades are observed within OPXV-PG-06, which correspond to the recently described MPXV Clade I and Clade II [4], with isolates from the 2022 global outbreak placed in Clade II (Fig 1). The phylogroup most closely related to MPXV is OPXV-PG-05 which contains all Vaccinia virus isolates, as well rabbitpox, buffalopox and horsepox viruses. The next closest virus was a so-called cowpox virus which was isolated from a cat [32] and does not cluster within any of the phylogroups. Other “cowpox” viruses have been isolated from a diverse set of host animals other than cows, such as humans, cats, alpaca and rodents (S1 Table). These viruses do not form a monophyletic clade but rather are seen as 4 distinct clades PG-02, PG-03, PG-04 and PG-07, each containing a mixture of both human and non-human derived isolates (Fig 1). The raccoonpox, volepox and skunkpox viruses were placed together in OPXV-PG-10 and showed much longer branch lengths making them the most distant members within the Orthopoxvirus genus (Fig 1). Analysis of pan-phylogroup proteomes formed by combining protein sequences from each isolate within a phylogroup, revealed common and distinct orthogroups present in various phylogroups (S2 Fig). A total of 162 orthogroups were present across the Orthopoxvirus genus as well as in the Centapox outgroup, and 14 orthogroups were present in the Orthopoxvirus genus but absent from the Centapox genus. This analysis also showed 36 orthogroups that were present only in the pan-MPXV proteome. However, none of these orthogroups were present across all MPXV isolates, precluding their use as MPXV specific biomarkers. Analysis of source of Orthopoxviruses from which they were isolated (S1 Table) revealed that viruses isolated from human were distributed throughout the tree (red asterisk in Fig 1) and interspersed with closely related isolates from various animal hosts. This distribution pattern indicates the dramatic zoonotic potential and disease risk of these viruses, highlighting the need for a pan-Orthopoxvirus detection assay.

**Fig 1.**
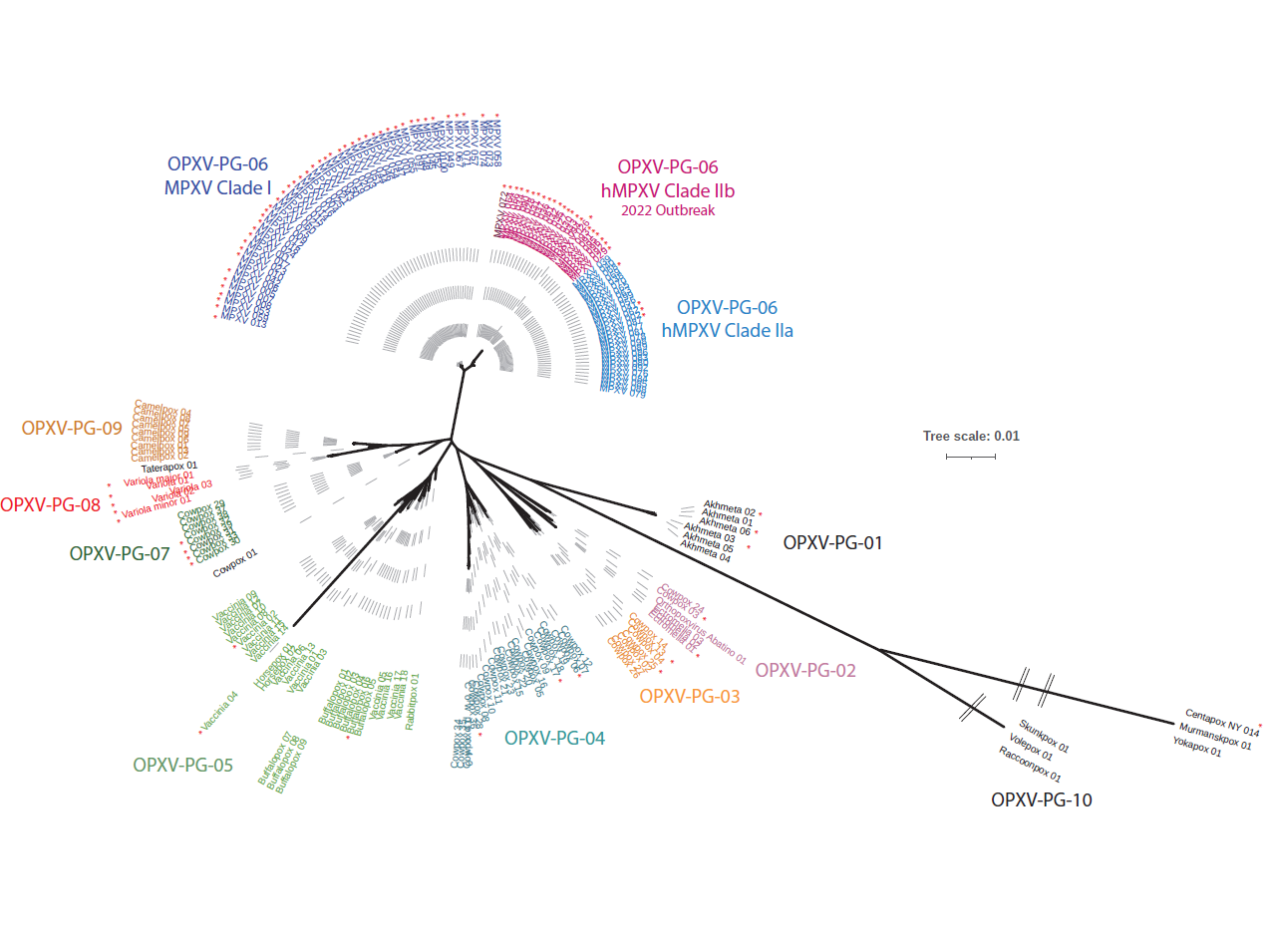
Phylogenomic tree of Orthopoxviruses. An unrooted phylogenomic tree of 200 Orthopoxviruses with genomes available in NCBI, including 100 genomes from various MPXV was constructed based on 44 single copy orthologs, with a combined supermatrix sequence length of 33,425 nucleotides. The Centapoxviruses were used as an outgroup. The substitution model GTR+F+I+G4 was found to be the best fit for the sequences. The phylogroups are color coded and indicated by corresponding labels. The virus isolates derived from infected humans are marked with red asterisks.

### Colorimetric LAMP assay development and sensitivity

A total of 14 sets of primers (S2 Table) targeting 8 different genes were tested in the colorimetric LAMP assays with fluorescent dye. Primers (Table 1) targeting the A4L (set 2) gene and the N1R (set 3) gene showed the best performance with the earliest signal detection and no background. The optimal LAMP reaction temperature was found to be 65°C with 40mM GuHCl included as an additive to speed up the reaction [33].

The sensitivity of the colorimetric A4L and N1R LAMP assays was first evaluated using a serial dilution of synthetic Mpox synthetic DNA. As shown in Fig 2A, synthetic DNA can be detected at 62.5 copies/µL in all triplicate reactions with either A4L or N1R LAMP primers. At 31.25 copies/µL, 8 out of 9 reactions (A4L LAMP) or 7 out of 9 reactions (N1R LAMP) showed a positive color change. As expected, the LAMP reaction containing both sets of LAMP primers improved the sensitivity and enabled the detection of all 9 reactions at 31.25 copies/µL.

**Fig 2.**
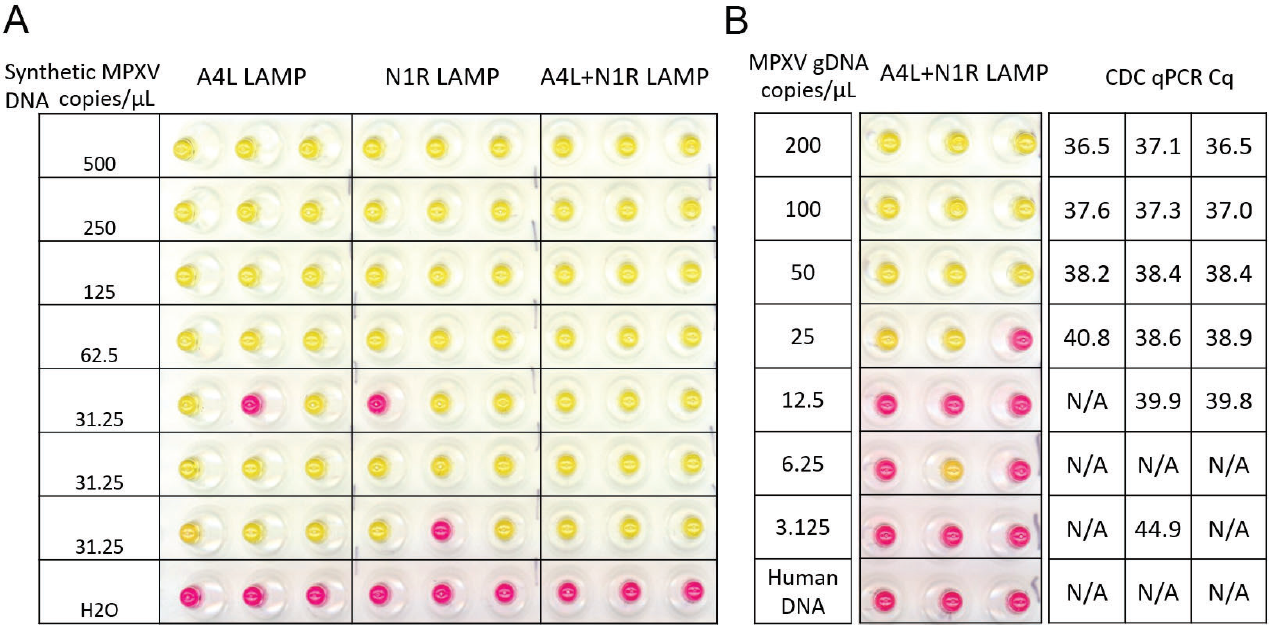
A4L and N1R LAMP assays can detect MPXV DNA. (A) MPXV synthetic DNA was diluted from 500 to 31.25 copies/µL in 0.1X TE buffer containing 1 ng/µL human DNA. 4 µL of DNA was tested in 20 µL LAMP reactions with either A4L primer set or N1R primer set or both. Scanned images of the post-amplification plate showing the colorimetric (pink = negative, yellow = positive) readouts are shown. (B) MPXV genomic DNA was diluted from 200 to 3.125 copies/µL and 4 µL of DNA was tested in colorimetric LAMP or CDC qPCR assays. The scanned image of the post-LAMP amplification plate and Cq number of CDC qPCR are shown. No amplification is denoted N/A. All reactions were performed in triplicates.

A direct comparison of the A4L+N1R colorimetric LAMP assay to the CDC Non-variola Orthopoxvirus Generic Real-Time PCR Test was performed using MPXV genomic DNA. As shown in Fig 2B, all replicates tested positive in the LAMP assay down to 50 copies/µL with relatively late Cq values (>38) in the CDC qPCR assay. At 25 copies/µL, 2 out of 3 reactions showed a positive color change in the LAMP assay, whereas an average Cq value of 39.4 was observed in the CDC qPCR assay. Therefore, the LAMP assay showed comparable sensitivity to the current gold standard qPCR assay, while being easier to perform.

### Specificity of A4L and N1R LAMP primer sets

The A4L gene is located in the more conserved central region of the viral genome and encodes the precursor of the essential major virion core protein p4b within the Orthopoxvirus. When the target DNA region of A4L LAMP was used to blast against all Monkeypox virus genomes in the NCBI database (on 3/31/2023), 4999 of the 5000 sequences are identical to the query sequence except for 1 sequence (OQ411312.1) due to one nucleotide N. The A4L gene sequence was also found to be almost identical in the majority of Orthopoxviruses with 1 to 3 nucleotide differences in the LAMP targeting region (S3 Fig). The most common difference is an A to C substitution which is located between the LB and B2 primer region and is not targeted by any primer, therefore it should not impact the performance of the LAMP reaction. The effect of nucleotide differences located within the LAMP primer binding sites such as single nucleotide differences located in either the F2 or F1c primer region (S3 Fig) were tested with genomic DNA from CMLV or VACV, which harbor these substitutions. As shown in Fig 3, these differences did not impact the ability of the A4L LAMP to detect the CMLV and VACV genomic DNAs, as successful amplifications were observed with Tt values within 10 minutes and a positive color change from pink to yellow (Fig 3) consistent with previous report on LAMP tolerance of sequence variation [34]. A blast search of the A4L target outside of the Orthopoxvirus genus found the Yoka poxvirus (Centapoxvirus genus within the Poxviridae family) as the closest hit with 79% sequence identity to the MPXV A4L gene (S3 Fig). Within the LAMP primer regions, 46 mismatched nucleotides were found, making it unlikely that the LAMP assay would amplify the Yoka poxvirus or other non-Orthopoxvirus. Taken together, these results indicate that the highly conserved A4L gene can be used as a pan-Orthopoxvirus biomarker in LAMP assays.

**Fig 3.**
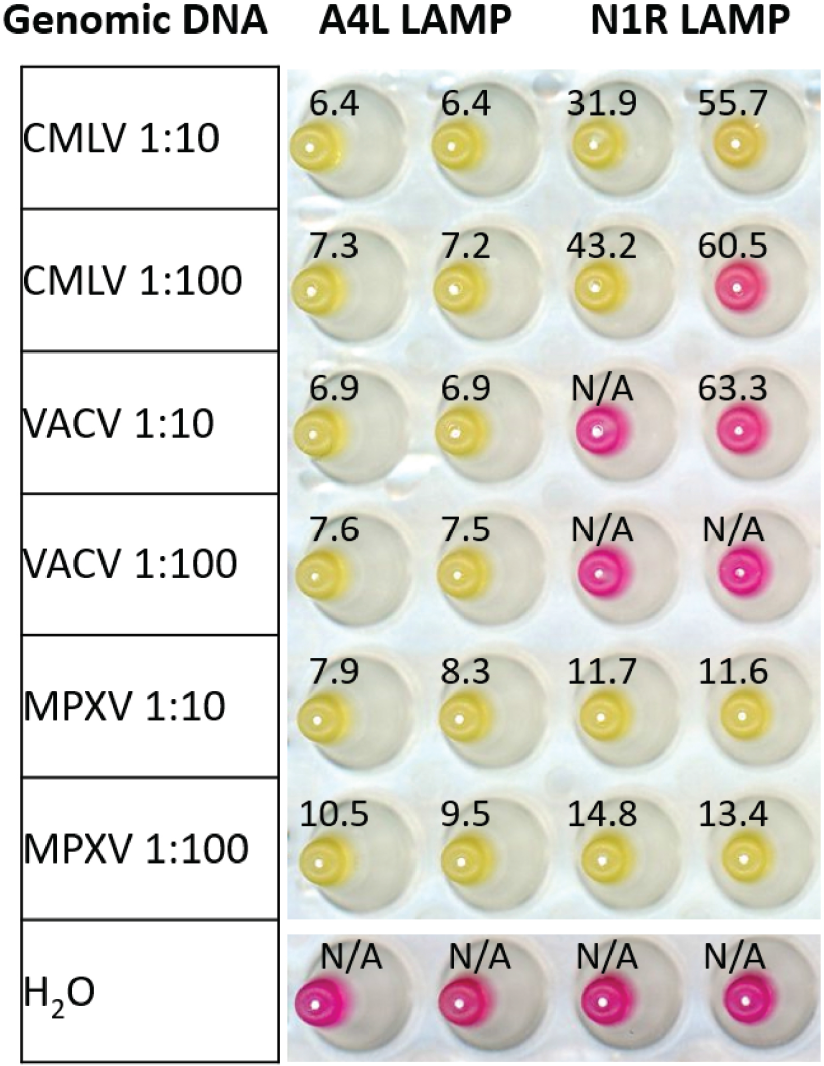
A4L and N1R LAMP assays on genomic DNA from various Orthopoxvirus. Genomic DNA from Camelpox virus (CMLV), Vaccinia virus (VACV), and Mpox virus (MPXV) diluted 10 or 100-fold were tested in A4L and N1R colorimetric LAMP assays containing 1µM SYTO™ 9 fluorescent dye. Real-time fluorescence and colorimetric (pink = negative, yellow = positive) readouts were obtained for each sample. The scanned image of the post-amplification plate is shown with overlaid Tt values defined as the time (min) to reach threshold fluorescence values during the amplification. No amplification is denoted N/A. Experiments were performed with two replicates.

The N1R gene is located towards the termini of the genome and encodes a Poxvirus Bcl-1-like protein, predicted to be involved in evading the host’s innate immune response. It is highly conserved among different MPXV isolates with 99.4% (4970/5000 on 3/31/2023) sequences in the NCBI database showing 100% identity to the query N1R LAMP target sequence while the remaining 0.6% (30/5000) sequences contain a single nucleotide difference located either in LF (A to G) or LB (C to T) primer binding region. No N1R homologs were found outside of the Orthopoxvirus genus based on the NCBI Blast search. Within Orthopoxvirus, the gene is more variable with multiple nucleotide differences (Fig 4 and S4 Fig). The effect of these differences was tested using genomic DNA from CMLV and VACV. CMLV N1R gene contains multiple nucleotide mismatches in the primer binding regions, including 3 in F3, 2 in B2, and 1 in LF. While 100-fold diluted MPXV DNA can be detected with a Tt value of 14 minutes, only 1 out of 2 reactions showed a positive result for CMLV DNA at that dilution, and with a significant delay of the amplification as indicated by a higher Tt value of 43 minutes (Fig 3) reflecting decreased amplification efficiency from mismatched nucleotides. No positive amplification was observed for VACV DNA because the corresponding sequence of this genomic DNA (NC_006998) lacks the N1R gene, different from other VACV isolates which contain the N1R gene with a similar number of mutations as CMLV (S4 Fig). Taken together, this N1R assay showed some selectivity towards MPXV compared with CMLV, it is not specific for MPXV.

**Fig 4.**
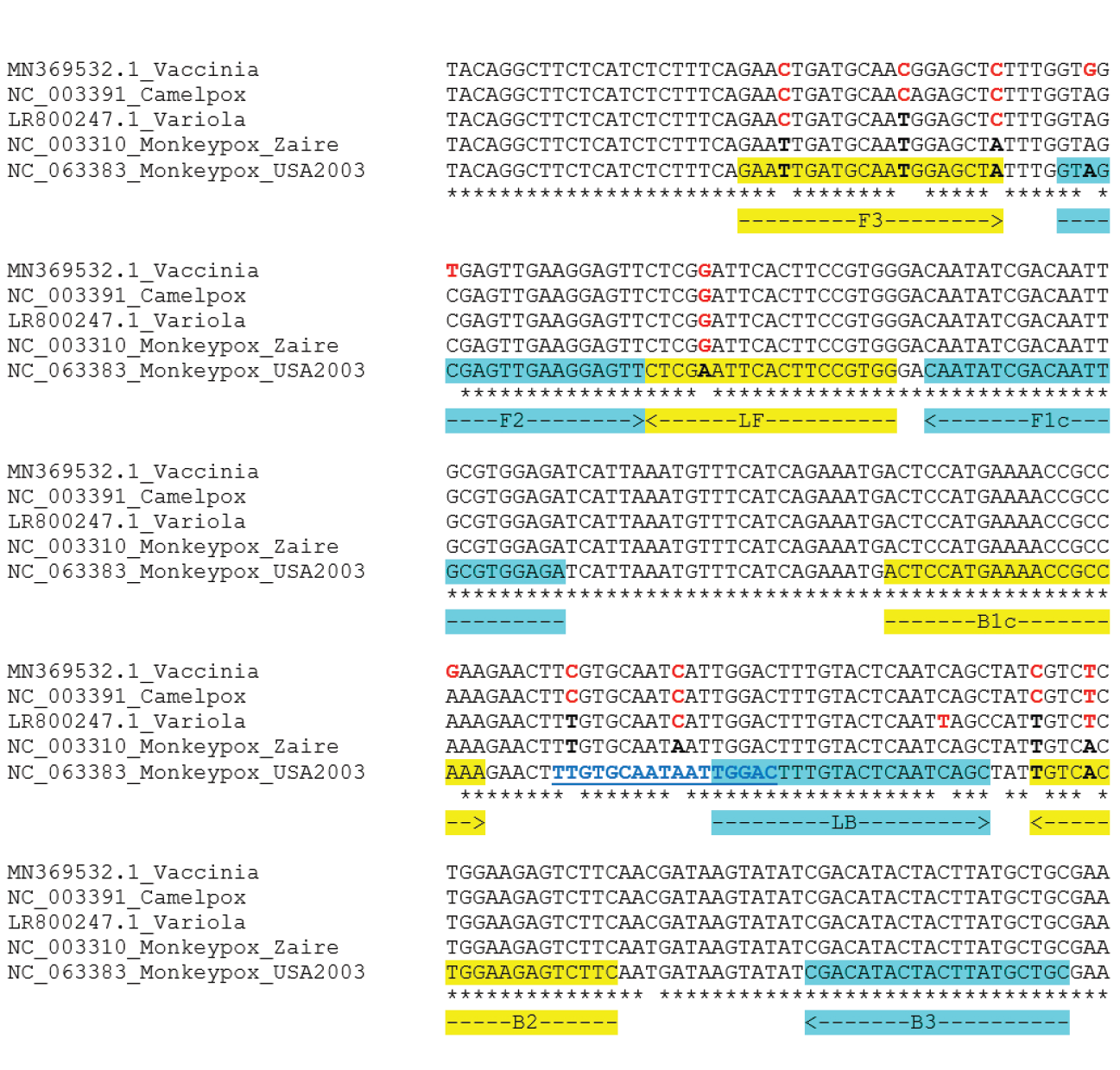
Alignment of N1R LAMP target sequence from various Orthopoxvirus. N1R target sequences were downloaded from NCBI and aligned with ClustalW. Nucleotides different from the MPXV sequence are indicated in bold red. Primer regions are highlighted in yellow or cyan. Nucleotide sequences used for the detection probe are shown in bold blue and underlined.

### Probe-based LAMP is specific for MPXV

Fluorescent detection probes with locked nucleic acid (LNA) bases have been used to distinguish SARS-CoV-2 variants in LAMP assays [35]. A probe targeting the N1R region between B1c and LB was designed to take advantage of the 2 nucleotide differences between CMLV and MPXV (Fig 4). When the probe was used in N1R LAMP reactions, amplification curves were observed only in the presence of MPXV genomic DNA, whereas no signal was detected with CMLV or VACV genomic DNA (Fig 5A), indicating the probe can distinguish DNA templates with 2 nucleotide difference in the probe binding region. While most of the N1R homologs were found to contain several nucleotide differences in the probe binding region, an exception was found in some variants of the Variola virus, for example in the genome of LR800247.1 (Fig 4) which contains only 1 nucleotide mismatch in the probe region. Due to the unavailability of Variola viral DNA for testing, a synthetic gBlock based on the MPXV backbone was designed to include all 6 nucleotide changes within the LAMP targeted region to mimic the variola N1R gene (S Text). N1R gBlocks for MPXV and CMLV were also synthesized and tested (Fig 5B). When a range of 10 to 10,000,000 copies of MPXV gBlocks was tested, 100 copies were reliably detected. In contrast, even at the highest copy number of 10,000,000, CMLV N1R gBlock could not be detected. For the variola mimic gBlock, very low amplification signals were observed, however, they were below the baseline fluorescence threshold and therefore scored negative. These experiments demonstrated that the probe-based LAMP assay can be used for specific detection of MPXV. To determine the sensitivity, a serial dilution of MPXV genomic DNA was tested, and reliable detection down to 62.5 copies/µL was observed around 20 minutes, whereas at 31.25 copies/µL, 8 of 9 reactions scored positive (Fig 5 C-D). Overall, this probe-based LAMP assay is both specific and sensitive. Representative amplification curves from 1 replicate are shown. (C) MPXV genomic DNA was serial diluted over a range of 500 to 31.25 copies/µL and tested in triplicate. (The Tt values measuring the incubation time in minutes to reach the fluorescent threshold were calculated from the Cq number and shown in (D).

**Fig 5.**
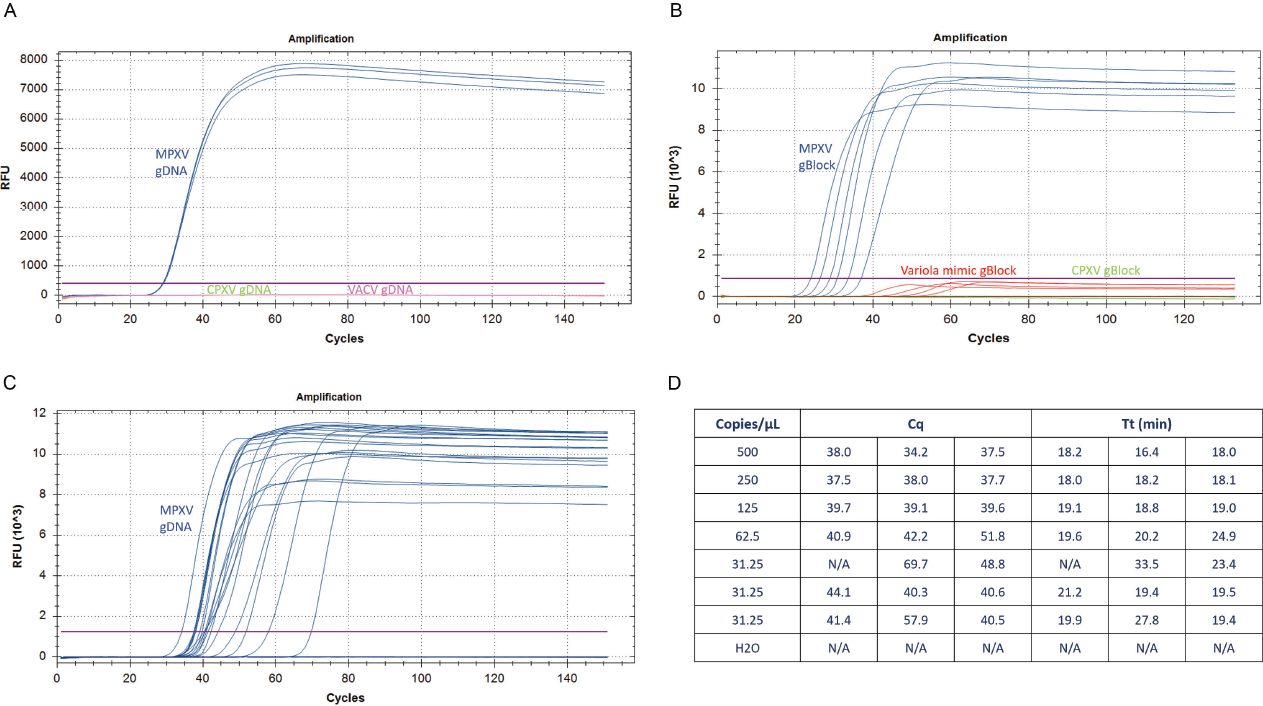
Specificity and sensitivity of probe-based LAMP assay. Probe-based LAMP reactions were performed in a Bio-Rad Opus qPCR machine with various DNA templates. The fluorescent amplification curves generated from the fluorescent probe are shown. (A) Genomic DNA from MPXV (blue), CMLV (green), and VACV (purple) were diluted 10-fold in 0.1X TE and tested in triplicate. (B) A 10-fold dilution series N1R gBlocks ranging from 5,000,000 to 5 copies/µL from MPXV (blue), CMLV (green), and the Variola mimic (red) were tested in duplicate.

### An extraction-free method for rapid detection of MPXV in clinical samples

Most molecular diagnostic protocols for MPXV detection involve DNA extraction from lesion material, such as lesion fluid on a dry swab. However, DNA extraction is a time-consuming and expensive process. An extraction-free colorimetric LAMP method has been successfully applied for SARS-CoV-2 detection in clinical samples [20]. This approach was evaluated and found to work well for MPXV detection directly from swab eluate when compared with the standard CDC qPCR method using purified DNA. After the eluates of lesion swabs from 3 patients were mixed with sample prep buffer and heated at 95°C for 5min, all three treated crude generated positive color change in the colorimetric LAMP assay (Fig 6). Consistent with the viral titer demonstrated by the qPCR assay, as little as 0.0625 µL of the eluate from the high viral containing Swab 1 is sufficient to generate a positive result in the colorimetric LAMP assay, while 2 µL eluate from the low viral containing Swab 3 was needed. Therefore, the extraction-free LAMP method displayed comparable performance to qPCR on purified DNA.

**Fig 6.**
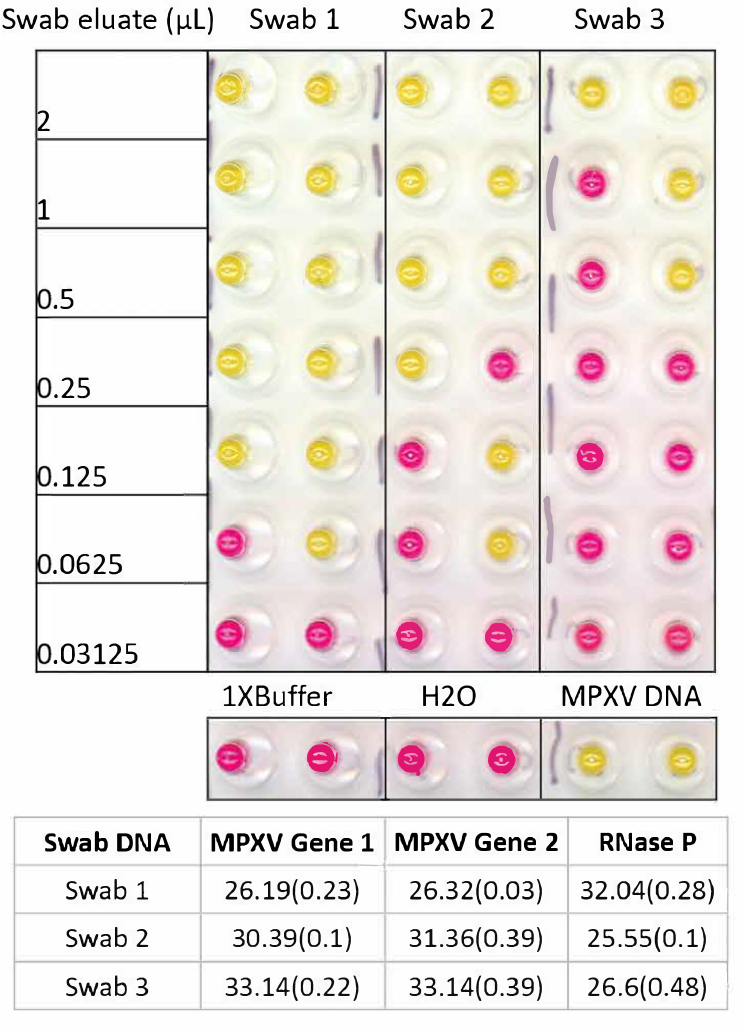
Direct virus detection of clinical swab eluate. Elute from the swab was mixed with an equal volume of 2X SLB buffer and heated at 95°C for 5min, then cooled to 4°C. For each LAMP reaction, 4µL of swab or serially diluted swab in 1X SLB was used. Water or 1XSLB buffer was used as the negative control, and MPXV Twist DNA was used as a positive control. The experiment was done in duplicate.

## Discussion

Members of the genus Orthopoxvirus are highly prevalent, zoonotic viruses that infect many mammals, and are particularly relevant to human health. While smallpox has been eradicated, the recent global outbreak in 2022 of Mpox caused by MPXV underscores the constant threat from these viruses and highlights the need for diagnostic tools for continued surveillance. The design and development of such assays require an understanding of the relationships between various Orthopoxviruses. Here we have utilized ∼200 genome sequences available in public databases to construct a comprehensive phylogenomic tree covering all known Orthopoxviruses. This analysis showed the presence of 10 distinct clades, denoted as phylogroups OPXV-PG-01 to OPXV-PG-10. No clear correspondence between the viruses within a phylogroup and their host species was found. For example, the so-called cowpox virus has been isolated from diverse animals, and these isolates were placed in multiple phylogroups, consistent with previous reports [32]. This demonstrates the importance of using genetic sequence and phylogenies for informative nomenclature and accurate classification for Orthopoxviruses, rather than naming them after the host species or their phenotypic properties [32]. Interestingly, various Orthopoxviruses which have been isolated from human subjects do not cluster together in a particular phylogroup but are distributed across the tree, illustrating that viruses from any of the phylogroups could be a threat to human health and agents of global outbreaks [36, 37]. A pan-Orthopoxvirus assay would be useful for monitoring animal reservoirs as well as new outbreaks in humans, while a species-specific viral test would be required for accurate diagnosis and treatment during a particular outbreak.

MPXV has been responsible for multiple outbreaks in recent decades. There are numerous ways to detect MPXV, including the detection of viral particles, virus-specific antigens, and many PCR-based methods [38]. The goals of the present study were to develop an isothermal, simple, rapid, and robust workflow for both pan-Orthopoxvirus and Mpox-specific diagnosis. In resource-limited settings, where access to thermocyclers and a stable electricity supply is challenging, LAMP-based assays are attractive. Several isothermal amplification-based assays, including LAMP and recombinase polymerase amplification, have been developed for diagnosing MPXV infection [39, 40] and used turbidimeters or gel electrophoresis instruments for readout, whereas our colorimetric LAMP test provides a simple visual readout.

One of the CDC’s clade-specific assays targets the tumor necrosis factor (TNF) receptor gene (G2R) [41, 42] which is located within the terminal inverted-repeat region and contains many single nucleotide polymorphisms and insertions/deletions specific for Clade II. However, in the recent outbreak, new deletions in the G2R target have negatively impacted the performance of the test resulting in dropouts. This led to a CDC advisory on September 2^nd^, 2022 [43], urging caution in the interpretation of negative results returned from the G2R assay if clinical suspicion for Mpox was high. The rapid accumulation of mutations in the 2022 MPXV isolates [44] necessitates the selection of more conserved targets. Our Orthopoxvirus colorimetric assay included two targets (A4L and N1R), the high conservation of the A4L gene in Orthopoxvirus and the high conservation of N1R within MPXV, combined with the tolerance of mismatches in LAMP reactions [34], are expected to prevent false negative results. The A4L LAMP region was conserved less in the skunkpox, raccoonpox, and volepox viruses with 19 to 30 mismatches in the primer binding region. These viruses form a distinct phylogroup with a very long branch in the phylogenomic tree indicating they are most distantly related to other Orthopoxviruses.

To achieve specific MPXV detection, a hybridization detection probe was designed and included in the N1R LAMP assay. The probe-based detection method requires no modification to the LAMP primers and can be used for the detection of single-nucleotide polymorphisms and small sequence changes [35]. The LAMP probe in our assay contains a 5′ end fluorophore and a 3’ end dark quencher. The probe only generates a fluorescent signal upon binding to N1R amplicons due to the increased structural rigidity of dsDNA and subsequent separation of the fluorophore and quencher. To enable using a short probe with high Tm, Locked Nucleic Acid (LNA) bases were incorporated into the probe, which is important to achieve specificity. The probe was designed to target the loop region of the LAMP amplicons to provide greater availability for probe annealing and less frequent displacement by the polymerase and showed little inhibition of the probe to the LAMP reaction. In our assay, a real-time PCR instrument was used for probe-based fluorescence detection. However, in the field, portable isothermal fluorimeters, endpoint plate readers, simple illumination instruments, or lateral flow can potentially be used for signal detection. In a recently published colorimetric LAMP assay targeting two different genes namely A27L and F3L, high specificity was claimed based on *in silico* analysis [45]. Using the published primer sets for A27L and F3L and their recommended reaction temperature, we found both CMLV and VACV DNA were detected (S5 Fig) indicating the assays lack specificity.

Most Mpox diagnostic assays in use, including the CDC-recommended assays, utilize purified DNA in qPCR reactions. In a rapidly expanding viral outbreak, testing can be limited by the time and cost of nucleic acid extraction steps, as witnessed during the SARS-CoV-2 pandemic. In our study, we used a simple extraction-free method involving a sample lysis buffer and heat treatment to release the DNA for detection directly from swab eluates. The heat treatment also inactivates the virus for quick and safe handling [46]. Detection of MPXV in saliva has been demonstrated to have a sensitivity of 88% compared to skin lesions [47]. Importantly, a saliva-based PCR test identified asymptomatic cases [48] before the appearance of rash or lesion, indicating that using a saliva test could help detect Mpox earlier in the time course of the illness. Earlier identification would likely reduce transmission. We expect our extraction-free colorimetric LAMP method could be used to detect MPXV in saliva as demonstrated previously for LAMP-based SARS-CoV-2 detection in saliva [20].

## Data Availability

All data produced in the present work are contained in the manuscript

## Acknowledgments

We thank Dr. Tom Evans for guidance and feedback on the manuscript.

## Supporting information

**S1 Fig. A mid-point rooted phylogenomic tree of 200 Orthopoxviruses, rendered in a rectangular layout.** The tree was constructed based on 44 single copy orthologs, with a combined supermatrix sequence length of 33,425 nucleotides. The Centapoxviruses were used as an outgroup. The substitution model GTR+F+I+G4 was found to be the best fit for the sequences. Branch lengths indicated on respective branches, which are not drawn to scale. are not to scale. The branches are colored by phylogroups. Bootstrap support values are shown at each node. The phylogroups are color-coded and indicated by corresponding labels.

S2 Fig. Analysis of orthogroups shared across various pan-phylogroup proteomes.

**S3 Fig. MPXV A4L sequence alignment.** MPXV A4L LAMP targeted sequence was used in the blast search excluding all the MPXV genomes to obtain all the homologs. (A). Multiple Sequence Alignment Viewer 1.23.0 (MSA viewer) of all the homologs within the Orthopoxvirus. Mismatched nucleotides were marked as red lines. (B). MPXV A4L LAMP targeted sequence alignment with homologs from CMLV, Variola virus, and VACV tested in the LAMP assay. Primer regions were highlighted with yellow or green color. Mismatched nucleotides were bold and colored red in related Orthopoxvirus. (C). A4L sequence alignment with the distantly related volepox, skunkpox, raccoonpox and yoka poxvirus, the closest related virus outside the Orthopoxvirus genus.

**S4 Fig. MPXV N1R sequence alignment.** MPXV N1E LAMP targeted sequence was used in the blast search excluding all the MPXV genomes to obtain all the homologs. Multiple Sequence Alignment Viewer 1.23.0 (MSA viewer) of all the homologs within the Orthopoxvirus. Mismatched nucleotides were marked as red lines.

**S5 Fig. Evaluation of A27L and F3L LAMP primers.** Synthetic human MPXV control DNA was diluted in human DNA (1ng/µL) from 1000 to 62.5 copies/µL and 4 µL of DNA was tested in 20 µL LAMP reactions with either A27L primer set or F3L primer set. LAMP reactions were performed at 63°C as the published reference described using NEB WarmStart Colorimetric LAMP mix. Scanned images of the post-amplification plate showing the colorimetric (pink/negative, yellow/positive) readouts were shown. Genomic DNA from camelpox virus (CMLV), vaccinia (VACV), and humans were also tested. Experiments were performed in triplicates.

S1 Table. List of Orthopoxviruses used for phylogenetic analysis.

S2 Table. List of 14 sets of LAMP primer sequences.

S3 Table. List of MPOX qPCR reagents and equipment.

S Text. gBlock sequence of variola N1R mimic and its alignment with N1R gene from MPXV and variola virus.

