## Supplementary material for "Development of an extraction-free LAMP method for the generic detection of Orthopoxvirus and for the specific detection of Mpox virus": S Text N1R__Mpox_Variola_Mimic.docx

```
Supplementary text: gBlock sequence of variola N1R mimic and its alignment with N1R gene from MPXV and variola


NC_063383_Monkeypox_USA2003      ATGGCCTCTCCTTGTGCCCAGTTCAGTCCCTGTCATTGCCACGCTACTAA
NC_003310_Monkeypox_Zaire        ATGGCCTTTCCTTGTGCCCAGTTCAGTCCCTGTCATTGCCACGCTACTAA
MPXV_N1R_Variola_Mimic_New       ATGGCCTTTCCTTGTGCCCAGTTCAGTCCCTGTCATTGCCACGCTACTAA
LR800247.1_Variola               ATGGCCTCTCCTTGTGCCCAGTTCAGTCCCTGTCATTGCCACGCTACTAA
                                 ******* ******************************************

NC_063383_Monkeypox_USA2003      GGACTCCCTGAATACCGTGACTGACGTCAGACATTGTCTGACTGAATACA
NC_003310_Monkeypox_Zaire        GGACTCCCTGAATACCGTGACTGACGTCAGACATTGTCTGACTGAATACA
MPXV_N1R_Variola_Mimic_New       GGACTCCCTGAATACCGTGACTGACGTCAGACATTGTCTGACTGAATACA
LR800247.1_Variola               GGACTCCCTGAATACCGTGGCCGACGTCAGACATTGTCTGACTGAATACA
                                 ******************* * ****************************

NC_063383_Monkeypox_USA2003      TCCTGTGGGTTTCTCATAGATGGACCCATAGAGAAAGCGCAGGGCCTCTC
NC_003310_Monkeypox_Zaire        TCCTGTGGGTTTCTCATAGATGGACCCATAGAGAAAGCGCAGGGCCTCTC
MPXV_N1R_Variola_Mimic_New       TCCTGTGGGTTTCTCATAGATGGACCCATAGAGAAAGCGCAGGGCCTCTC
LR800247.1_Variola               TCCTGTGGGTTTCTCATAGATGGACCCATAGAGAAAGCGCAGGGTCTCTC
                                 ******************************************** *****

NC_063383_Monkeypox_USA2003      TACAGGCTTCTCATCTCTTTCAGAATTGATGCAATGGAGCTATTTGGTAG
NC_003310_Monkeypox_Zaire        TACAGGCTTCTCATCTCTTTCAGAATTGATGCAATGGAGCTATTTGGTAG
MPXV_N1R_Variola_Mimic_New       TACAGGCTTCTCATCTCTTTCAGAACTGATGCAATGGAGCTCTTTGGTAG
LR800247.1_Variola               TACAGGCTTCTCATCTCTTTCAGAACTGATGCAATGGAGCTCTTTGGTAG
                                 ************************* *************** ********
                                                       ---------F3-------->    ----

NC_063383_Monkeypox_USA2003      CGAGTTGAAGGAGTTCTCGAATTCACTTCCGTGGGACAATATCGACAATT
NC_003310_Monkeypox_Zaire        CGAGTTGAAGGAGTTCTCGGATTCACTTCCGTGGGACAATATCGACAATT
MPXV_N1R_Variola_Mimic_New       CGAGTTGAAGGAGTTCTCGGATTCACTTCCGTGGGACAATATCGACAATT
LR800247.1_Variola               CGAGTTGAAGGAGTTCTCGGATTCACTTCCGTGGGACAATATCGACAATT
                                 ******************* ******************************
                                 ----F2--------><------LF----------  <-------F1c---

NC_063383_Monkeypox_USA2003      GCGTGGAGATCATTAAATGTTTCATCAGAAATGACTCCATGAAAACCGCC
NC_003310_Monkeypox_Zaire        GCGTGGAGATCATTAAATGTTTCATCAGAAATGACTCCATGAAAACCGCC
MPXV_N1R_Variola_Mimic_New       GCGTGGAGATCATTAAATGTTTCATCAGAAATGACTCCATGAAAACCGCC
LR800247.1_Variola               GCGTGGAGATCATTAAATGTTTCATCAGAAATGACTCCATGAAAACCGCC
                                 **************************************************
                                 ---------                        -------B1c-------

NC_063383_Monkeypox_USA2003      AAAGAACTTTGTGCAATAATTGGACTTTGTACTCAATCAGCTATTGTCAC
NC_003310_Monkeypox_Zaire        AAAGAACTTTGTGCAATAATTGGACTTTGTACTCAATCAGCTATTGTCAC
MPXV_N1R_Variola_Mimic_New       AAAGAACTTTGTGCAATCATTGGACTTTGTACTCAATTAGCTATTGTCTC
LR800247.1_Variola               AAAGAACTTTGTGCAATCATTGGACTTTGTACTCAATTAGCCATTGTCTC
                                 ***************** ******************* *** ****** *
                                 -->                 ---------LB--------->   <-----

NC_063383_Monkeypox_USA2003      TGGAAGAGTCTTCAATGATAAGTATATCGACATACTACTTATGCTGCGAA
NC_003310_Monkeypox_Zaire        TGGAAGAGTCTTCAATGATAAGTATATCGACATACTACTTATGCTGCGAA
MPXV_N1R_Variola_Mimic_New       TGGAAGAGTCTTCAATGATAAGTATATCGACATACTACTTATGCTGCGAA
LR800247.1_Variola               TGGAAGAGTCTTCAACGATAAGTATATCGACATACTACTTATGCTGCGAA
                                 *************** **********************************
                                 -----B2------              <-------B3----------

NC_063383_Monkeypox_USA2003      AGATTCTGAACGAGAACGACTATCTCACCCTCTTGGATCATATCCTCACT
NC_003310_Monkeypox_Zaire        AGATTCTGAATGAGAACGACTATCTCACCCTCTTGGATCATATCCTCACT
MPXV_N1R_Variola_Mimic_New       AGATTCTGAATGAGAACGACTATCTCACCCTCTTGGATCATATCCTCACT
LR800247.1_Variola               AGATTCTGAACGAGAACGACTATCTCACCCTCTTGGATCATATCCGCACT
                                 ********** ********************************** ****
```
