## Supplementary figures and images for "Development of an extraction-free LAMP method for the generic detection of Orthopoxvirus and for the specific detection of Mpox virus"

### S1 Fig.pdf

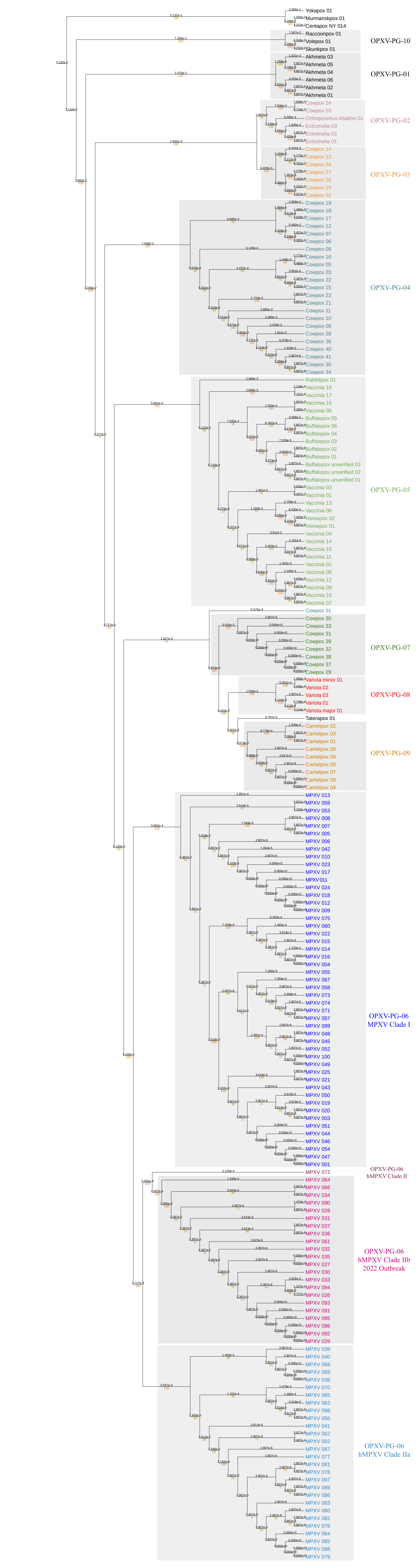

### S2 Fig.pdf

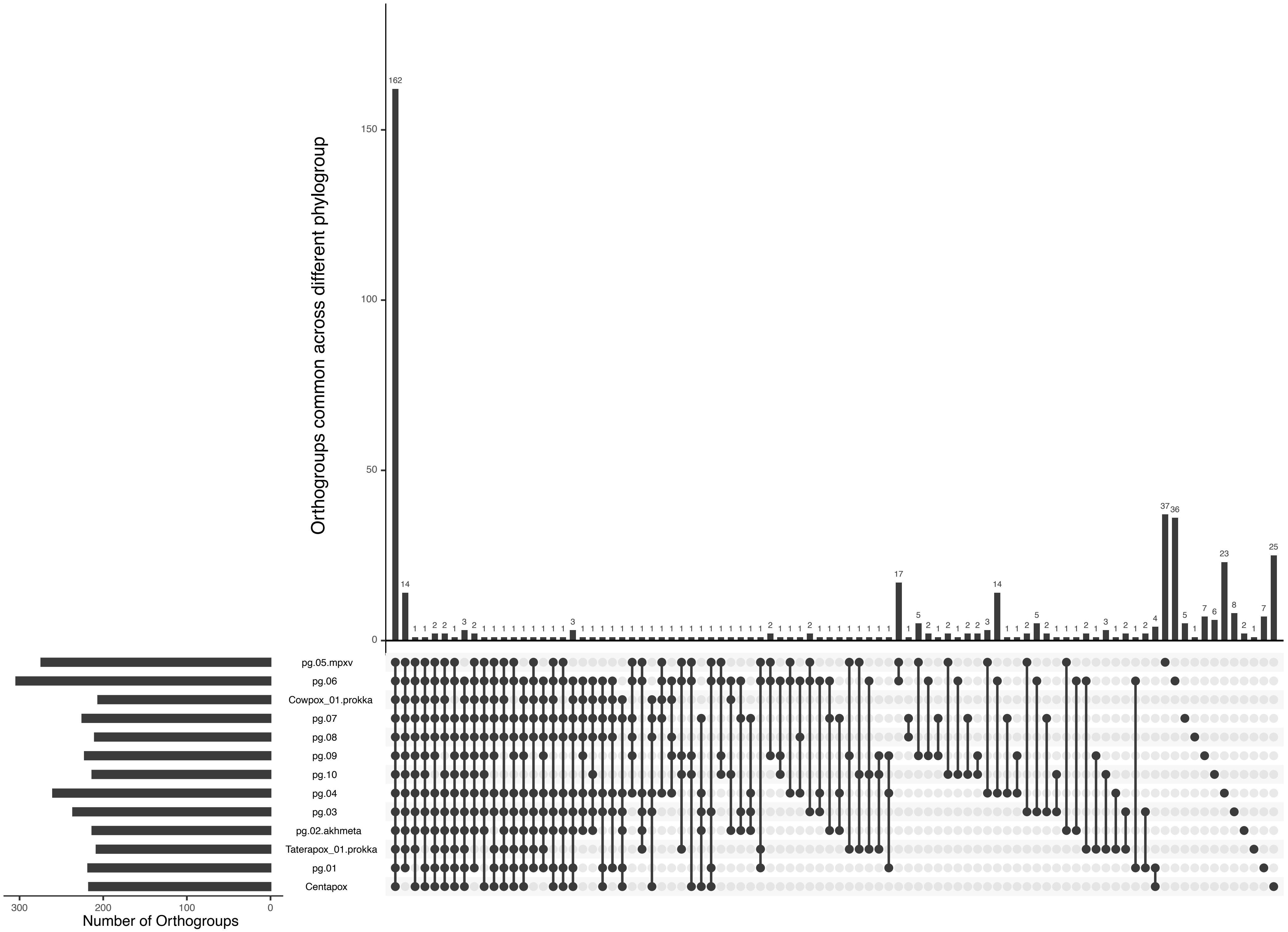

### S3 Fig.pdf

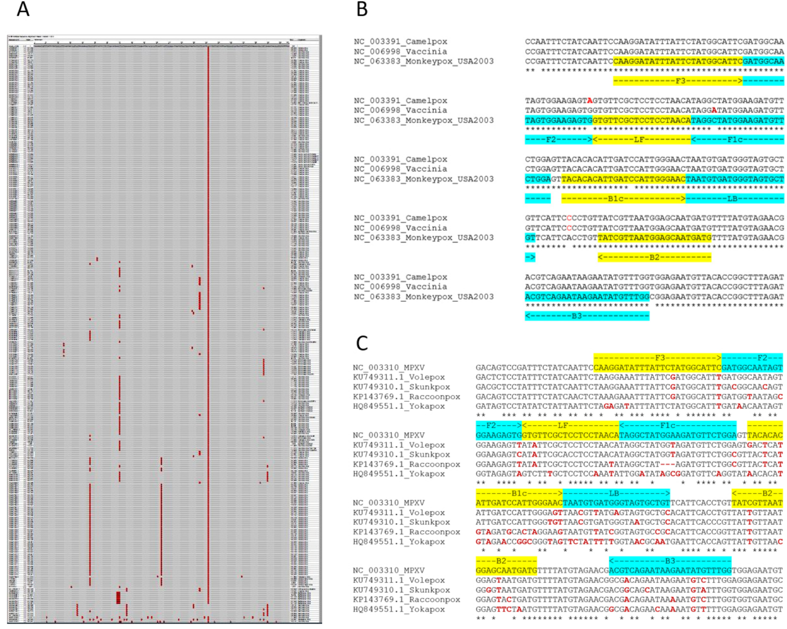

### S4 Fig N1R_LAMPproduct_Alignment_nonMPXV_Orthopox.pdf

NCBI Multiple Sequence Alignment Viewer, Version 1.22.0

| Sequence ID | Start | Alignment | End | Organism |
| --- | --- | --- | --- | --- |

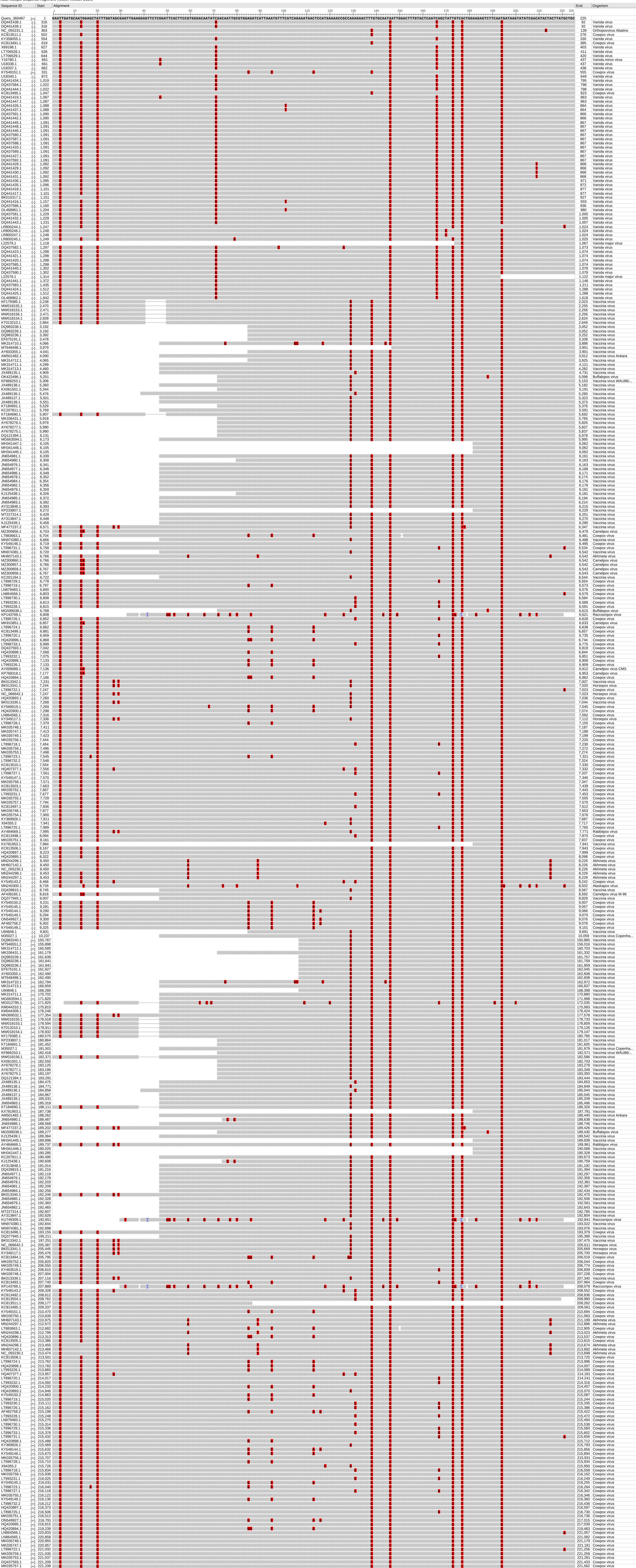
